## Supplementary material for "Protective effect of Mediterranean type glucose-6-phosphate dehydrogenase deficiency against Plasmodium vivax malaria": Code and data: Meta_Analysis_G6PDMed.pdf

### Gene-dose effect of G6PDd Med in the Pasthun ethnic group

James Watson

27/02/2019

```
knitr::opts_chunk$set(cache = TRUE, cache.comments = FALSE,
                      include = TRUE, echo = TRUE,
                      fig.width = 9, fig.height = 9,
                      fig.pos = 'H',
                      dev = 'png', dpi = 300)
library(rstan)
```

```
## Loading required package: StanHeaders

## Loading required package: ggplot2

## rstan (Version 2.21.2, GitRev: 2e1f913d3ca3)

## For execution on a local, multicore CPU with excess RAM we recommend calling
## options(mc.cores = parallel::detectCores()).
## To avoid recompilation of unchanged Stan programs, we recommend calling
## rstan_options(auto_write = TRUE)
```

```
library(StanHeaders)
library(bayesplot)
```

```
## This is bayesplot version 1.7.2

## - Online documentation and vignettes at mc-stan.org/bayesplot

## - bayesplot theme set to bayesplot::theme_default()

##    * Does _not_ affect other ggplot2 plots

##    * See ?bayesplot_theme_set for details on theme setting
```

```
library(ggplot2)
library(gridExtra)
library(HardyWeinberg)
```

```
## Loading required package: mice

##
## Attaching package: 'mice'

## The following objects are masked from 'package:base':
##
##     cbind, rbind

## Loading required package: Rsolnp
```

```
version
```

```
## function (pkg = "mice")
## {
```

```
##     lib <- dirname(system.file(package = pkg))
##     d <- packageDescription(pkg)
##     return(paste(d$Package, d$Version, d$Date, lib))
## }
## <bytecode: 0x7ff2b9c7bd08>
## <environment: namespace:mice>
```

```r
sessionInfo()
```

```
## R version 4.0.2 (2020-06-22)
## Platform: x86_64-apple-darwin17.0 (64-bit)
## Running under: macOS Catalina 10.15.2
##
## Matrix products: default
## BLAS:   /Library/Frameworks/R.framework/Versions/4.0/Resources/lib/libRblas.dylib
## LAPACK: /Library/Frameworks/R.framework/Versions/4.0/Resources/lib/libRlapack.dylib
##
## locale:
## [1] en_US.UTF-8/en_US.UTF-8/en_US.UTF-8/C/en_US.UTF-8/en_US.UTF-8
##
## attached base packages:
## [1] stats     graphics  grDevices utils     datasets  methods   base
##
## other attached packages:
## [1] HardyWeinberg_1.6.8  Rsolnp_1.16          mice_3.10.0
## [4] gridExtra_2.3        bayesplot_1.7.2      rstan_2.21.2
## [7] ggplot2_3.3.2        StanHeaders_2.21.0-5
##
## loaded via a namespace (and not attached):
##  [1] tidyselect_1.1.0   xfun_0.15          purrr_0.3.4        lattice_0.20-41
##  [5] V8_3.2.0           colorspace_1.4-1   vctrs_0.3.2        generics_0.0.2
##  [9] htmltools_0.5.0    stats4_4.0.2       loo_2.3.1          yaml_2.2.1
## [13] rlang_0.4.7        pkgbuild_1.1.0     pillar_1.4.6       glue_1.4.1
## [17] withr_2.2.0        matrixStats_0.56.0 lifecycle_0.2.0    plyr_1.8.6
## [21] stringr_1.4.0      munsell_0.5.0      gtable_0.3.0       codetools_0.2-16
## [25] evaluate_0.14      inline_0.3.15      knitr_1.29         callr_3.4.3
## [29] ps_1.3.3           curl_4.3           parallel_4.0.2     fansi_0.4.1
## [33] broom_0.7.0        Rcpp_1.0.5.1       scales_1.1.1       backports_1.1.8
## [37] RcppParallel_5.0.2 jsonlite_1.7.0     truncnorm_1.0-8    digest_0.6.25
## [41] stringi_1.4.6      processx_3.4.3     dplyr_1.0.0        grid_4.0.2
## [45] cli_2.0.2          tools_4.0.2        magrittr_1.5       tibble_3.0.3
## [49] crayon_1.3.4       tidyr_1.1.0        pkgconfig_2.0.3    ellipsis_0.3.1
## [53] prettyunits_1.1.1  ggridges_0.5.2     assertthat_0.2.1   rmarkdown_2.3
## [57] R6_2.4.1           compiler_4.0.2
```

### Data

##### parasite counts

```r
para_dat = readr::read_csv('Data/Parasite_counts.csv')
```

```
## Parsed with column specification:
## cols(
```

```
##   Hb = col_double(),
##   `Age in years` = col_double(),
##   `Parasite/uL` = col_double(),
##   `Gamet/uL` = col_double(),
##   Parasitaemia = col_double(),
##   PCR = col_character()
## )
```

```r
para_dat$PCR[grep('homozygous', para_dat$PCR,ignore.case = T)] = 0
para_dat$PCR[grep('hetero', para_dat$PCR,ignore.case = T)] = 1
para_dat$PCR[grep('WT', para_dat$PCR, ignore.case = T)] = 2
para_dat$PCR = as.numeric(para_dat$PCR)
table(para_dat$PCR)
```

```
##
##   0   1   2
##   6  25 570
```

```r
par(las = 1)
hist(log10(para_dat$Parasitaemia))
```

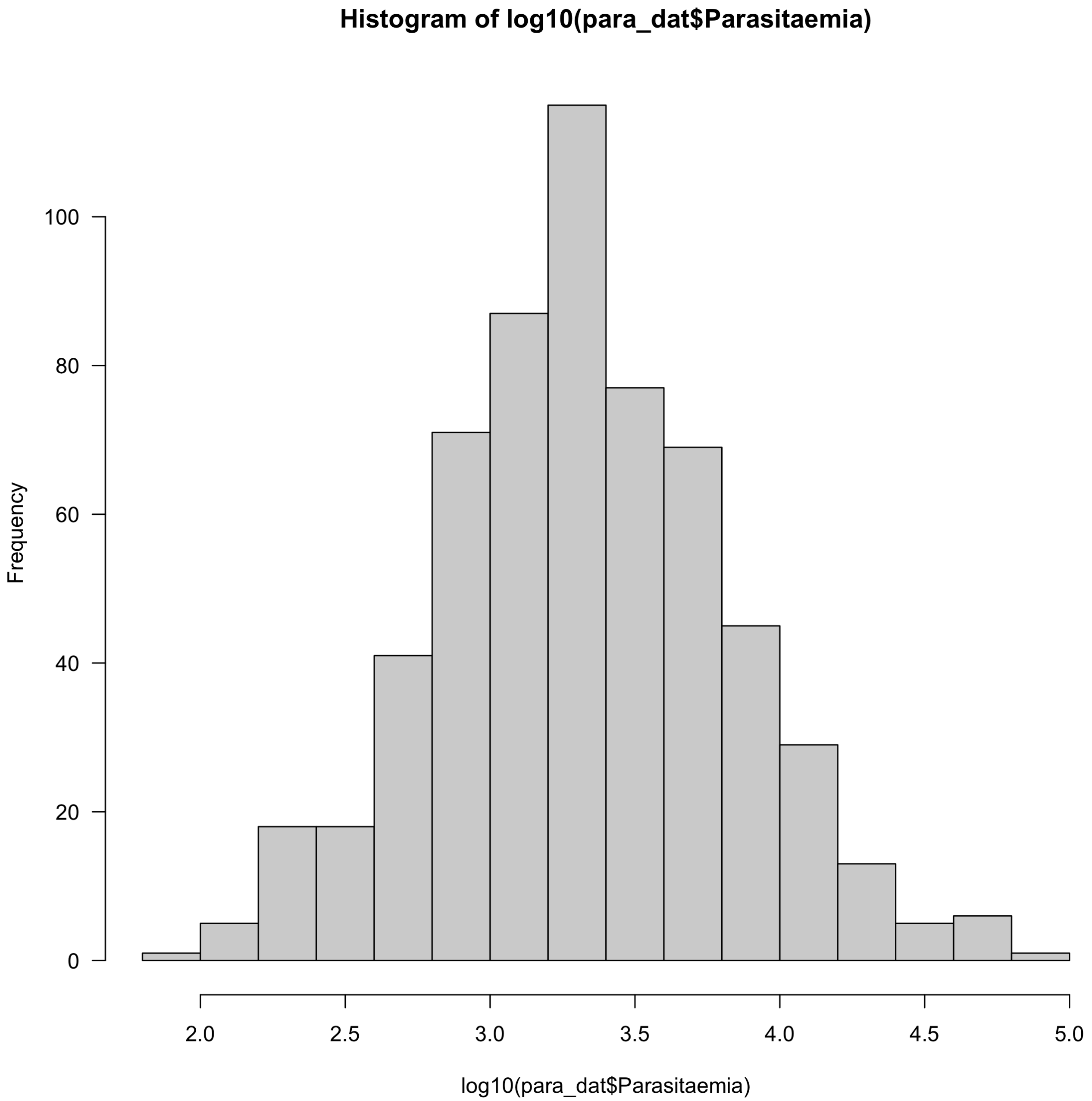


```
boxplot(log10(para_dat$Parasitaemia) ~ para_dat$PCR, xaxt='n',
        ylab='Log 10 parasitaemia', xlab='', yaxt='n')
axis(2, at = log10(c(100,300,1000,3000,10000,30000)),
     labels =c(100,300,1000,3000,10000,30000))
axis(1, at =1:3, labels = c('Homo/hemizygous','Heterozygous','WT'))
```

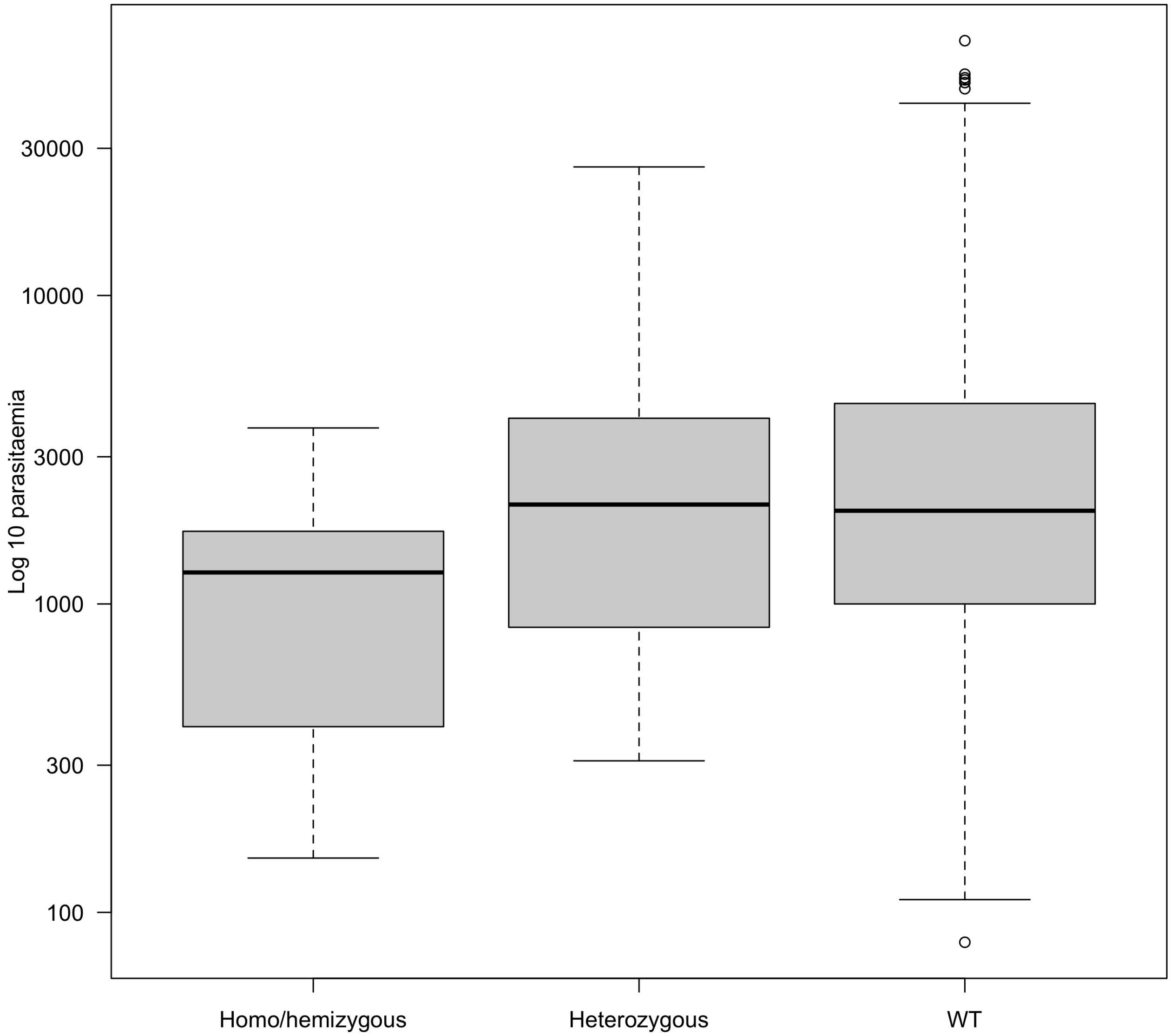


```r
mod = glm(PCR ~ log10para, family='binomial', weights = rep(2, nrow(para_dat)),
    data = data.frame(PCR = para_dat$PCR/2,
                      log10para=log10(para_dat$Parasitaemia)))
summary(mod)
```

```
##
## Call:
## glm(formula = PCR ~ log10para, family = "binomial", data = data.frame(PCR = para_dat$PCR/2,
##     log10para = log10(para_dat$Parasitaemia)), weights = rep(2,
##     nrow(para_dat)))
##
## Deviance Residuals:
##     Min       1Q   Median       3Q      Max
## -3.8097   0.3172   0.3474   0.3751   0.4900
##
## Coefficients:
##             Estimate Std. Error z value Pr(>|z|)
## (Intercept)   1.8483     1.1017   1.678   0.0934 .
## log10para     0.4910     0.3398   1.445   0.1484
```

```
## ---
## Signif. codes:  0 '***' 0.001 '**' 0.01 '*' 0.05 '.' 0.1 ' ' 1
##
## (Dispersion parameter for binomial family taken to be 1)
##
##     Null deviance: 261.12  on 600  degrees of freedom
## Residual deviance: 259.00  on 599  degrees of freedom
## AIC: 297.66
##
## Number of Fisher Scoring iterations: 6
```

```
wilcox.test(para_dat$Parasitaemia[para_dat$PCR==0],
            para_dat$Parasitaemia[para_dat$PCR>=1])
```

```
##
##  Wilcoxon rank sum test with continuity correction
##
## data:  para_dat$Parasitaemia[para_dat$PCR == 0] and para_dat$Parasitaemia[para_dat$PCR >= 1]
## W = 1117, p-value = 0.1147
## alternative hypothesis: true location shift is not equal to 0
```

##### Hb exclusion criteria

```
pcr_before = readr::read_csv('Data/PCR_data_before_cutoff.csv')
```

```
## Parsed with column specification:
## cols(
##   Sex = col_character(),
##   Hb = col_double(),
##   PCR = col_character()
## )
```

```
pcr_after = readr::read_csv('Data/PCR_data_after_cutoff.csv')
```

```
## Parsed with column specification:
## cols(
##   Sex = col_character(),
##   Hb = col_double(),
##   PCR = col_character()
## )
```

```
table(pcr_after$Sex, pcr_after$PCR)
```

```
##
##     Heterozygous Homozygous  WT
##   F            7          0 103
##   M            2          3 186
```

```
table(pcr_before$Sex, pcr_before$PCR)
```

```
##
##          Heterozygous (563C>C/T) Homozygous (563C>T) unable to amplify  WT
##   Female                      23                   3                 2 314
##   Male                         0                   2                 0 115
```

```r
pcr_before$Hb = as.numeric(pcr_before$Hb)
pcr_after$Hb = as.numeric(pcr_after$Hb)

pcr_before$PCR[grep('homozygous', pcr_before$PCR,ignore.case = T)] = 0
pcr_before$PCR[grep('hetero', pcr_before$PCR,ignore.case = T)] = 1
pcr_before$PCR[grep('WT', pcr_before$PCR, ignore.case = T)] = 2
pcr_before$PCR[grep('unable', pcr_before$PCR, ignore.case = T)] = NA

pcr_after$PCR[grep('homozygous', pcr_after$PCR,ignore.case = T)] = 0
pcr_after$PCR[grep('hetero', pcr_after$PCR,ignore.case = T)] = 1
pcr_after$PCR[grep('WT', pcr_after$PCR, ignore.case = T)] = 2

table(pcr_before$Sex, pcr_before$PCR)
```

```
##
##             0   1   2
##   Female    3  23 314
##   Male      2   0 115
```

```r
table(pcr_after$Sex, pcr_after$PCR)
```

```
##
##       0   1   2
##   F   0   7 103
##   M   3   2 186
```

```r
pcr_after$Sex[pcr_after$Sex=='M' & pcr_after$PCR==1]='F'
par(las= 1, mfrow=c(2,2))
hist(pcr_before$Hb, xlim = c(5, 20), breaks = seq(5,20,by=1),
     main = 'Before',xlab='Hb (g/dL)')
abline(v = 8, col='red',lwd=3)
hist(pcr_after$Hb, xlim = c(5, 20), breaks = seq(5,20,by=1),
     main = 'After',xlab='Hb (g/dL)')
abline(v = 8, col='red',lwd=3)

boxplot(pcr_before$Hb ~ pcr_before$PCR, ylab='Hb (g/dL)',xlab = '',xaxt='n')
axis(1, at =1:3, labels = c('Homo/hemizygous','Heterozygous','WT'))
boxplot(pcr_after$Hb ~ pcr_after$PCR, ylab='Hb (g/dL)',xlab = '',xaxt='n')
axis(1, at = 1:3, labels = c('Homo/hemizygous','Heterozygous','WT'))
```

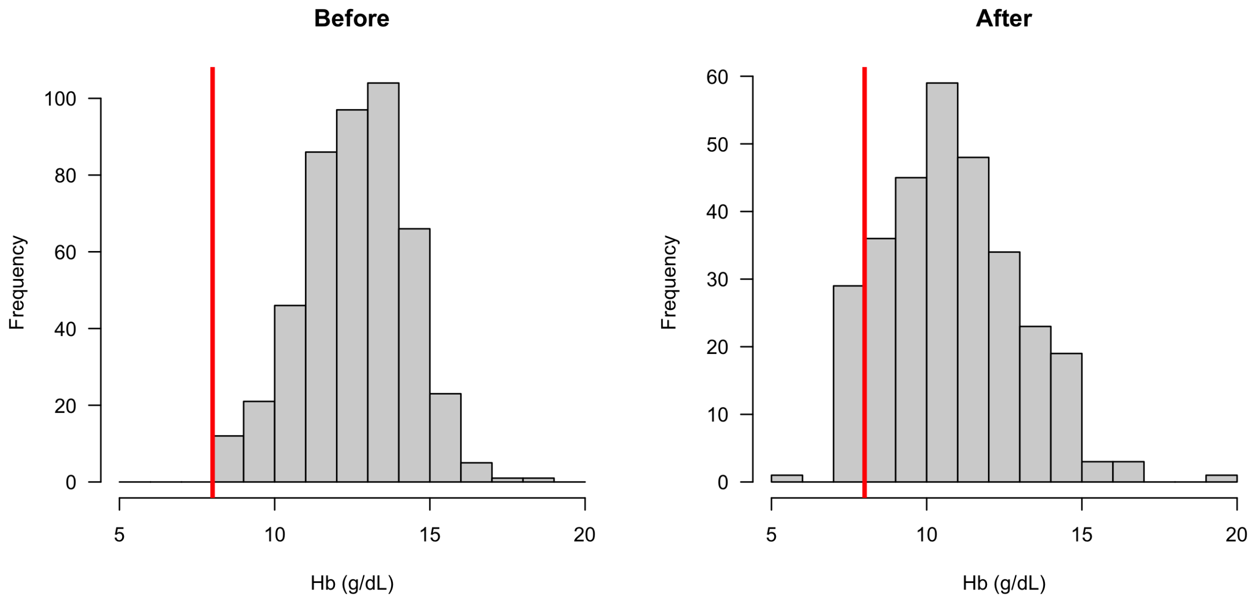


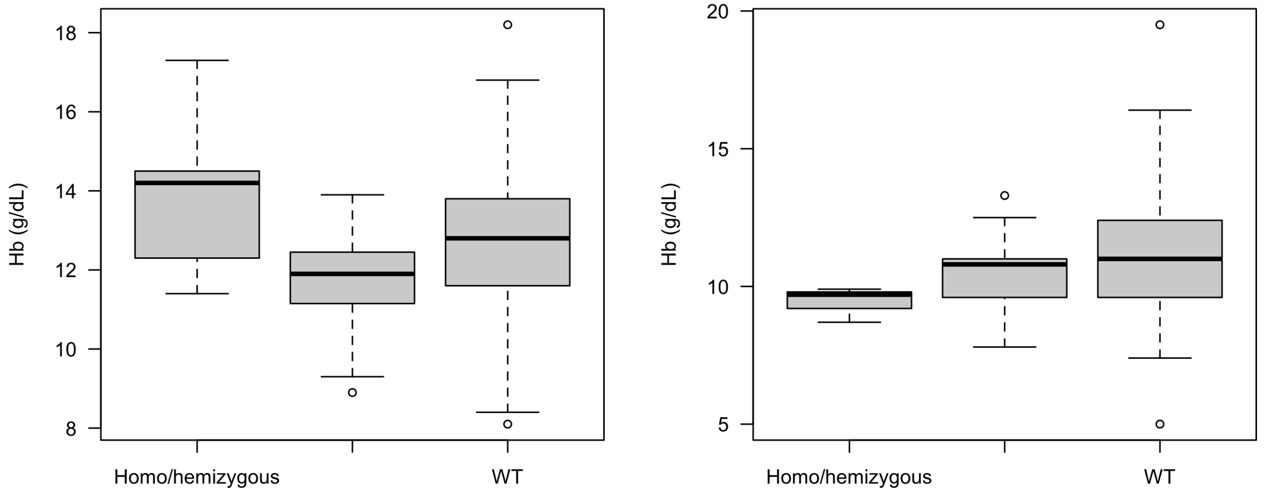


```
mod = glm(PCR ~ Hb, family='binomial', weights = rep(2, nrow(pcr_before)),
    data = data.frame(PCR = as.numeric(pcr_before$PCR)/2,
                      Hb=pcr_before$Hb))
summary(mod)
```

```
##
## Call:
## glm(formula = PCR ~ Hb, family = "binomial", data = data.frame(PCR = as.numeric(pcr_before$PCR)/2,
##     Hb = pcr_before$Hb), weights = rep(2, nrow(pcr_before)))
##
## Deviance Residuals:
##     Min       1Q   Median       3Q      Max
## -3.8970   0.3541   0.3761   0.3994   0.4780
##
## Coefficients:
##             Estimate Std. Error z value Pr(>|z|)
```

```
## (Intercept)   2.0065     1.3222   1.518    0.129
## Hb            0.1022     0.1059   0.965    0.334
##
## (Dispersion parameter for binomial family taken to be 1)
##
##     Null deviance: 220.23  on 456  degrees of freedom
## Residual deviance: 219.31  on 455  degrees of freedom
##   (5 observations deleted due to missingness)
## AIC: 255.19
##
## Number of Fisher Scoring iterations: 6
```

```
mod = glm(PCR ~ Hb, family='binomial', weights = rep(2, nrow(pcr_after)),
    data = data.frame(PCR = as.numeric(pcr_after$PCR)/2,
                      Hb=pcr_after$Hb))
summary(mod)
```

```
##
## Call:
## glm(formula = PCR ~ Hb, family = "binomial", data = data.frame(PCR = as.numeric(pcr_after$PCR)/2,
##     Hb = pcr_after$Hb), weights = rep(2, nrow(pcr_after)))
##
## Deviance Residuals:
##     Min       1Q   Median       3Q      Max
## -3.7679   0.2503   0.3037   0.3489   0.5730
##
## Coefficients:
##             Estimate Std. Error z value Pr(>|z|)
## (Intercept)   1.3754     1.3816   0.996    0.319
## Hb            0.2166     0.1336   1.622    0.105
##
## (Dispersion parameter for binomial family taken to be 1)
##
##     Null deviance: 115.44  on 300  degrees of freedom
## Residual deviance: 112.62  on 299  degrees of freedom
## AIC: 129.1
##
## Number of Fisher Scoring iterations: 6
```

```
awab_dat = rbind(pcr_after, pcr_before)
awab_dat$Sex[awab_dat$Sex %in% c('F','Female')] = 'F'
awab_dat$Sex[awab_dat$Sex %in% c('M','Male')] = 'M'
table(awab_dat$Sex)
```

```
##
##   F   M
## 456 307
```

```
awab_dat = awab_dat[!is.na(awab_dat$PCR), ]
```

```
# Awab study
Awabdata = list(Nmales_controls_hemi = 28,
                Nmales_controls_total = 314+28,
                Nmales_vivaxcases_hemi = 5,
                Nmales_vivaxcases_total = 5+299,
                Nfemales_controls_homo = 2,
```

```
                Nfemales_controls_het = 50,
                Nfemales_controls_total = 305+50+2,
                Nfemales_vivaxcases_homo = 3,
                Nfemales_vivaxcases_het = 32,
                Nfemales_vivaxcases_total = 425+32+3)

# Combined data
# Leslie + Bouma + Awab
Combineddata = list(Nmales_controls_hemi=31+25+Awabdata$Nmales_controls_hemi,
                    Nmales_controls_total=285+31+214+25+Awabdata$Nmales_controls_total,
                    Nmales_vivaxcases_hemi=2+0+Awabdata$Nmales_vivaxcases_hemi,
                    Nmales_vivaxcases_total=155+2+Awabdata$Nmales_vivaxcases_total,
                    Nfemales_controls_homo=2+0+Awabdata$Nfemales_controls_homo,
                    Nfemales_controls_het=26+0+Awabdata$Nfemales_controls_het,
                    Nfemales_controls_total=126+26+2+Awabdata$Nfemales_controls_total,
                    Nfemales_vivaxcases_homo=0+Awabdata$Nfemales_vivaxcases_homo,
                    Nfemales_vivaxcases_het=6+Awabdata$Nfemales_vivaxcases_het,
                    Nfemales_vivaxcases_total=72+6+Awabdata$Nfemales_vivaxcases_total)

datAwab = data.frame(sex = c(rep('male',
                                 Awabdata$Nmales_controls_total+
                                   Awabdata$Nmales_vivaxcases_total),
                             rep('female',
                                 Awabdata$Nfemales_controls_total+
                                   Awabdata$Nfemales_vivaxcases_total)),
                     G6PD = c(rep('hemi/homo',Awabdata$Nmales_controls_hemi),
                              rep('Normal',Awabdata$Nmales_controls_total-
                                    Awabdata$Nmales_controls_hemi),
                              rep('hemi/homo',Awabdata$Nmales_vivaxcases_hemi),
                              rep('Normal',Awabdata$Nmales_vivaxcases_total-
                                    Awabdata$Nmales_vivaxcases_hemi),
                              rep('hemi/homo',Awabdata$Nfemales_controls_homo),
                              rep('Het',Awabdata$Nfemales_controls_het),
                              rep('Normal',Awabdata$Nfemales_controls_total-
                                    Awabdata$Nfemales_controls_homo-
                                    Awabdata$Nfemales_controls_het),
                              rep('hemi/homo',Awabdata$Nfemales_vivaxcases_homo),
                              rep('Het',Awabdata$Nfemales_vivaxcases_het),
                              rep('Normal',Awabdata$Nfemales_vivaxcases_total-
                                    Awabdata$Nfemales_vivaxcases_homo-
                                    Awabdata$Nfemales_vivaxcases_het)),
                     Status = c(rep('Healthy',Awabdata$Nmales_controls_total),
                                rep('Vivax',Awabdata$Nmales_vivaxcases_total),
                                rep('Healthy',Awabdata$Nfemales_controls_total),
                                rep('Vivax',Awabdata$Nfemales_vivaxcases_total)))

datCombined = data.frame(sex = c(rep('male',
                                     Combineddata$Nmales_controls_total+
                                       Combineddata$Nmales_vivaxcases_total),
                                 rep('female',
                                     Combineddata$Nfemales_controls_total+
                                       Combineddata$Nfemales_vivaxcases_total)),
                         G6PD = c(rep('hemi/homo',Combineddata$Nmales_controls_hemi),
```

```
                              rep('Normal',Combineddata$Nmales_controls_total-
                                    Combineddata$Nmales_controls_hemi),
                              rep('hemi/homo',Combineddata$Nmales_vivaxcases_hemi),
                              rep('Normal',Combineddata$Nmales_vivaxcases_total-
                                    Combineddata$Nmales_vivaxcases_hemi),
                              rep('hemi/homo',Combineddata$Nfemales_controls_homo),
                              rep('Het',Combineddata$Nfemales_controls_het),
                              rep('Normal',Combineddata$Nfemales_controls_total-
                                    Combineddata$Nfemales_controls_homo-
                                    Combineddata$Nfemales_controls_het),
                              rep('hemi/homo',Combineddata$Nfemales_vivaxcases_homo),
                              rep('Het',Combineddata$Nfemales_vivaxcases_het),
                              rep('Normal',Combineddata$Nfemales_vivaxcases_total-
                                    Combineddata$Nfemales_vivaxcases_homo-
                                    Combineddata$Nfemales_vivaxcases_het)),
                        Status = c(rep('Healthy',Combineddata$Nmales_controls_total),
                               rep('Vivax',Combineddata$Nmales_vivaxcases_total),
                               rep('Healthy',Combineddata$Nfemales_controls_total),
                               rep('Vivax',Combineddata$Nfemales_vivaxcases_total)))
writeLines('In Awab\'s data, the breakdown is as follows:\n')
```

```
## In Awab's data, the breakdown is as follows:
```

```
knitr::kable(table(datAwab$sex,datAwab$G6PD,datAwab$Status))
```

| Var1 | Var2 | Var3 | Freq |
|---|---|---|---|
| female | hemi/homo | Healthy | 2 |
| male | hemi/homo | Healthy | 28 |
| female | Het | Healthy | 50 |
| male | Het | Healthy | 0 |
| female | Normal | Healthy | 305 |
| male | Normal | Healthy | 314 |
| female | hemi/homo | Vivax | 3 |
| male | hemi/homo | Vivax | 5 |
| female | Het | Vivax | 32 |
| male | Het | Vivax | 0 |
| female | Normal | Vivax | 425 |
| male | Normal | Vivax | 299 |

```
writeLines('In the combined data, the breakdown is as follows:\n')
```

```
## In the combined data, the breakdown is as follows:
```

```
knitr::kable(table(datCombined$sex,datCombined$G6PD,datCombined$Status))
```

| Var1 | Var2 | Var3 | Freq |
|---|---|---|---|
| female | hemi/homo | Healthy | 4 |
| male | hemi/homo | Healthy | 84 |
| female | Het | Healthy | 76 |
| male | Het | Healthy | 0 |
| female | Normal | Healthy | 431 |
| male | Normal | Healthy | 813 |
| female | hemi/homo | Vivax | 3 |

| Var1 | Var2 | Var3 | Freq |
|---|---|---|---|
| male | hemi/homo | Vivax | 7 |
| female | Het | Vivax | 38 |
| male | Het | Vivax | 0 |
| female | Normal | Vivax | 497 |
| male | Normal | Vivax | 454 |

### Testing Hardy-Weinberg

```
Awab_controls = c(A = Awabdata$Nmales_controls_hemi,
                  B = Awabdata$Nmales_controls_total-
                    Awabdata$Nmales_controls_hemi,
                  AA = Awabdata$Nfemales_controls_homo,
                  AB = Awabdata$Nfemales_controls_het,
                  BB = Awabdata$Nfemales_controls_total-
                    Awabdata$Nfemales_controls_homo-
                    Awabdata$Nfemales_controls_het )

All_controls = c(A = Combineddata$Nmales_controls_hemi,
                 B = Combineddata$Nmales_controls_total-
                   Combineddata$Nmales_controls_hemi,
                 AA = Combineddata$Nfemales_controls_homo,
                 AB = Combineddata$Nfemales_controls_het,
                 BB = Combineddata$Nfemales_controls_total-
                   Combineddata$Nfemales_controls_homo-
                   Combineddata$Nfemales_controls_het )

HWExact(Awab_controls,x.linked=TRUE,verbose=TRUE)
```

```
## Graffelman-Weir exact test for Hardy-Weinberg equilibrium on the X-chromosome
## using SELOME p-value
## Sample probability 0.02651387 p-value =  0.9436494
```

```
HWExact(All_controls,x.linked=TRUE,verbose=TRUE)
```

```
## Graffelman-Weir exact test for Hardy-Weinberg equilibrium on the X-chromosome
## using SELOME p-value
## Sample probability 0.008872498 p-value =  0.5968529
```

#### Stan model

The assumptions in the model:

- Hardy-Weinberg equilibrium holds in the Pashtun group (there is no evidence that this assumption does not hold in the controls)
- The protective effect is the same in hemizygous deficient males as it is in homozygous deficient females

```
# stan model code
G6PD_effect = "

data {
  int<lower=0> Nmales_controls_hemi;           // #males hemizygous deficient
```

```
  int<lower=0> Nmales_controls_total;          // #males controls total
  int<lower=0> Nfemales_controls_homo;         // #females homozygous deficient
  int<lower=0> Nfemales_controls_het;          // #females heterozygous deficient
  int<lower=0> Nfemales_controls_total;        // #females controls total
  int<lower=0> Nmales_vivaxcases_hemi;         // #males hemizygous deficient vivax
  int<lower=0> Nmales_vivaxcases_total;        // #males total vivax
  int<lower=0> Nfemales_vivaxcases_homo;       // #females homozygous deficient vivax
  int<lower=0> Nfemales_vivaxcases_het;        // #females heterozygous deficient vivax
  int<lower=0> Nfemales_vivaxcases_total;      // #females total vivax
  real p_1;                                    // Prior beta model of prevalence p
  real p_2;                                    // Prior beta model of prevalence p
}

transformed data {
  int female_controls[3];
  int female_cases[3];
  female_controls[1] = Nfemales_controls_homo;
  female_controls[2] = Nfemales_controls_het;
  female_controls[3] = Nfemales_controls_total-Nfemales_controls_het-Nfemales_controls_homo;
  female_cases[1] = Nfemales_vivaxcases_homo;
  female_cases[2] = Nfemales_vivaxcases_het;
  female_cases[3] = Nfemales_vivaxcases_total-Nfemales_vivaxcases_het-Nfemales_vivaxcases_homo;
}

parameters {
  real<lower=0,upper=1> p;      // allele frequency
  real<lower=0,upper=1> alpha;  // protective effect in fully deficients: hemi or homo-zygous
  real<lower=0,upper=1> beta;   // protective effect in heterozygous deficients
}

transformed parameters {
  // For ease of model interpretation (these are the quantities we are interested in)
  real one_minus_alpha = 1-alpha;
  real one_minus_beta = 1-beta;
  vector[3] theta_females_controls;
  vector[3] theta_females_cases;
  theta_females_controls[1] = p^2;
  theta_females_controls[2] = 2*p*(1-p);
  theta_females_controls[3] = 1 - 2*p*(1-p) - p^2;
  theta_females_cases[1] = alpha * p^2;
  theta_females_cases[2] = beta * 2*p*(1-p);
  theta_females_cases[3] = 1 - (beta*2*p*(1-p)) - (alpha*p^2);
}

model {
  // Prior
  p ~ beta(p_1, p_2);
  alpha ~ uniform(0,1);
  beta ~ uniform(0,1);

  // Multinomial likelihood for females and binomial likelihood for males
  // Healthy individuals - controls : gives estimate for p
  Nmales_controls_hemi ~ binomial(Nmales_controls_total, p);
```

```
  female_controls ~ multinomial(theta_females_controls);

  //Vivax individuals: gives estimate for alpha and beta
  Nmales_vivaxcases_hemi ~ binomial(Nmales_vivaxcases_total, alpha*p);
  female_cases ~ multinomial(theta_females_cases);

}
"
G6PD_effect_stan = stan_model(model_code = G6PD_effect)
```

```
# Awab's data
options(mc.cores = 4)
set.seed(7581)


writeLines(sprintf('In Awab data, we have %s controls (%s males and %s females) and %s vivax cases (%s m
                   Awabdata$Nmales_controls_total+Awabdata$Nfemales_controls_total,
                   Awabdata$Nmales_controls_total,Awabdata$Nfemales_controls_total,
                   Awabdata$Nmales_vivaxcases_total+Awabdata$Nfemales_vivaxcases_total,
                   Awabdata$Nmales_vivaxcases_total,Awabdata$Nfemales_vivaxcases_total))
```

```
## In Awab data, we have 699 controls (342 males and 357 females) and 764 vivax cases (304 males and 460
```

```
## Just Awab data
mod_awab=sampling(G6PD_effect_stan,
                  data=c(Awabdata, p_1 = 2, p_2 = 18),
                  iter = 10^6, thin = 2*10^2, chains=4)

thetas1=extract(mod_awab)
knitr::kable(round(100*sapply(thetas1, quantile, probs = c(0.025,.5,.975)),1)[,1:5])
```

| | p | alpha | beta | one_minus_alpha | one_minus_beta |
|---|---|---|---|---|---|
| 2.5% | 6.3 | 14.9 | 33.4 | 38.6 | 28.3 |
| 50% | 7.8 | 32.4 | 49.3 | 67.6 | 50.7 |
| 97.5% | 9.5 | 61.4 | 71.7 | 85.1 | 66.6 |

```
writeLines(sprintf('In the combined dataset, we have %s controls (%s males and %s females) and %s vivax
                   Combineddata$Nmales_controls_total+Combineddata$Nfemales_controls_total,
                   Combineddata$Nmales_controls_total,Combineddata$Nfemales_controls_total,
                   Combineddata$Nmales_vivaxcases_total+Combineddata$Nfemales_vivaxcases_total,
                   Combineddata$Nmales_vivaxcases_total,Combineddata$Nfemales_vivaxcases_total))
```

```
## In the combined dataset, we have 1408 controls (897 males and 511 females) and 999 vivax cases (461 m
```

```
# Meta-analysis: Awab+Leslie+Bouma
mod_combined=sampling(G6PD_effect_stan,
                      data=c(Combineddata, p_1 = 2, p_2 = 18),
                      iter = 10^6, thin = 2*10^2, chains=4)

thetas2=extract(mod_combined)
knitr::kable(round(100*sapply(thetas2, quantile, probs = c(0.025,.5,.975)),1)[,1:5])
```

| | p | alpha | beta | one_minus_alpha | one_minus_beta |
|---|---|---|---|---|---|
| 2.5% | 7.6 | 11.9 | 32.0 | 58.2 | 38.4 |

| | p | alpha | beta | one_minus_alpha | one_minus_beta |
|---|---|---|---|---|---|
| 50% | 8.8 | 23.9 | 45.0 | 76.1 | 55.0 |
| 97.5% | 10.1 | 41.8 | 61.6 | 88.1 | 68.0 |

Check model convergence

```
traceplot(mod_awab)
```

```
## 'pars' not specified. Showing first 10 parameters by default.
```

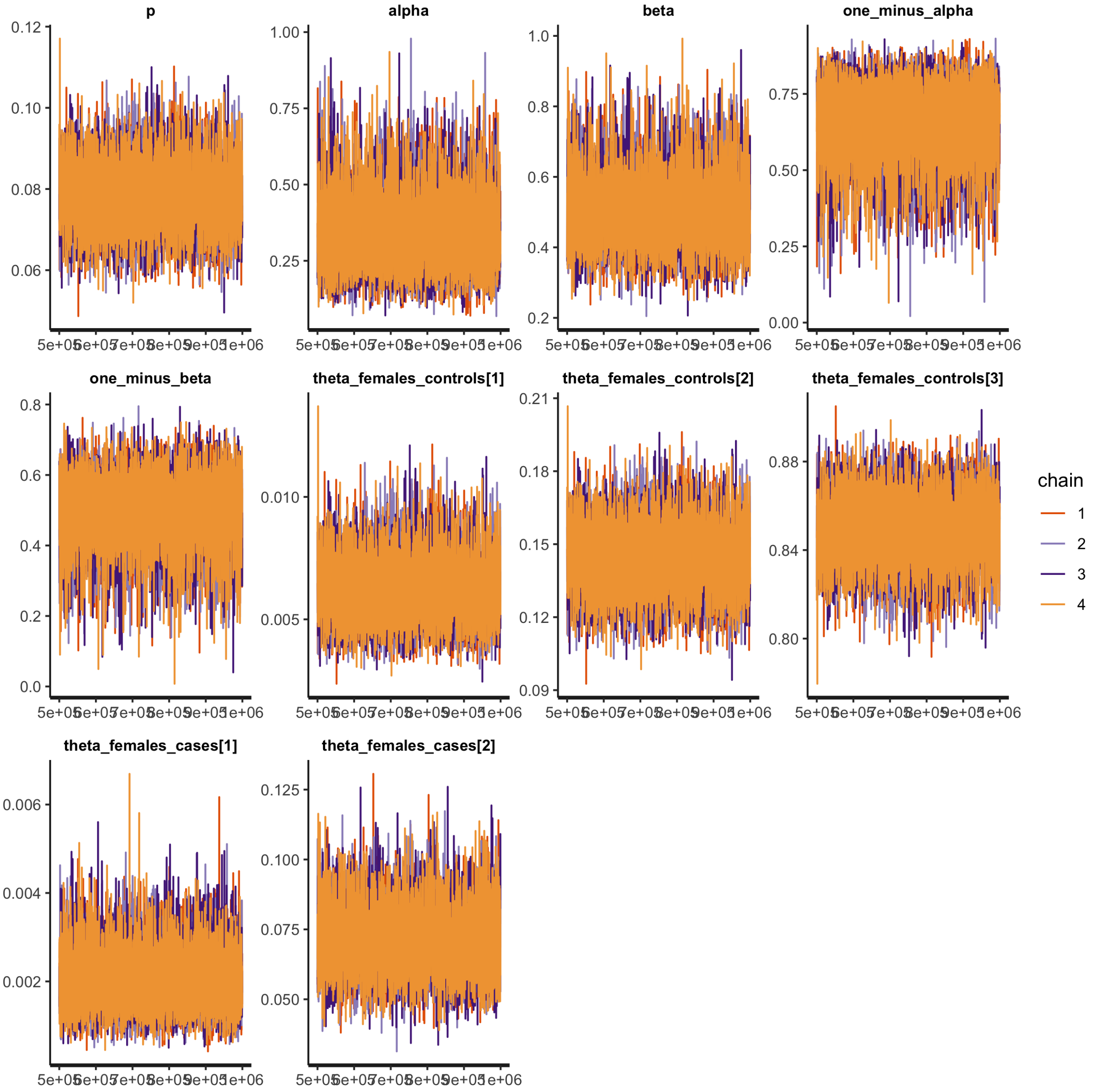


```
traceplot(mod_combined)
```

```
## 'pars' not specified. Showing first 10 parameters by default.
```

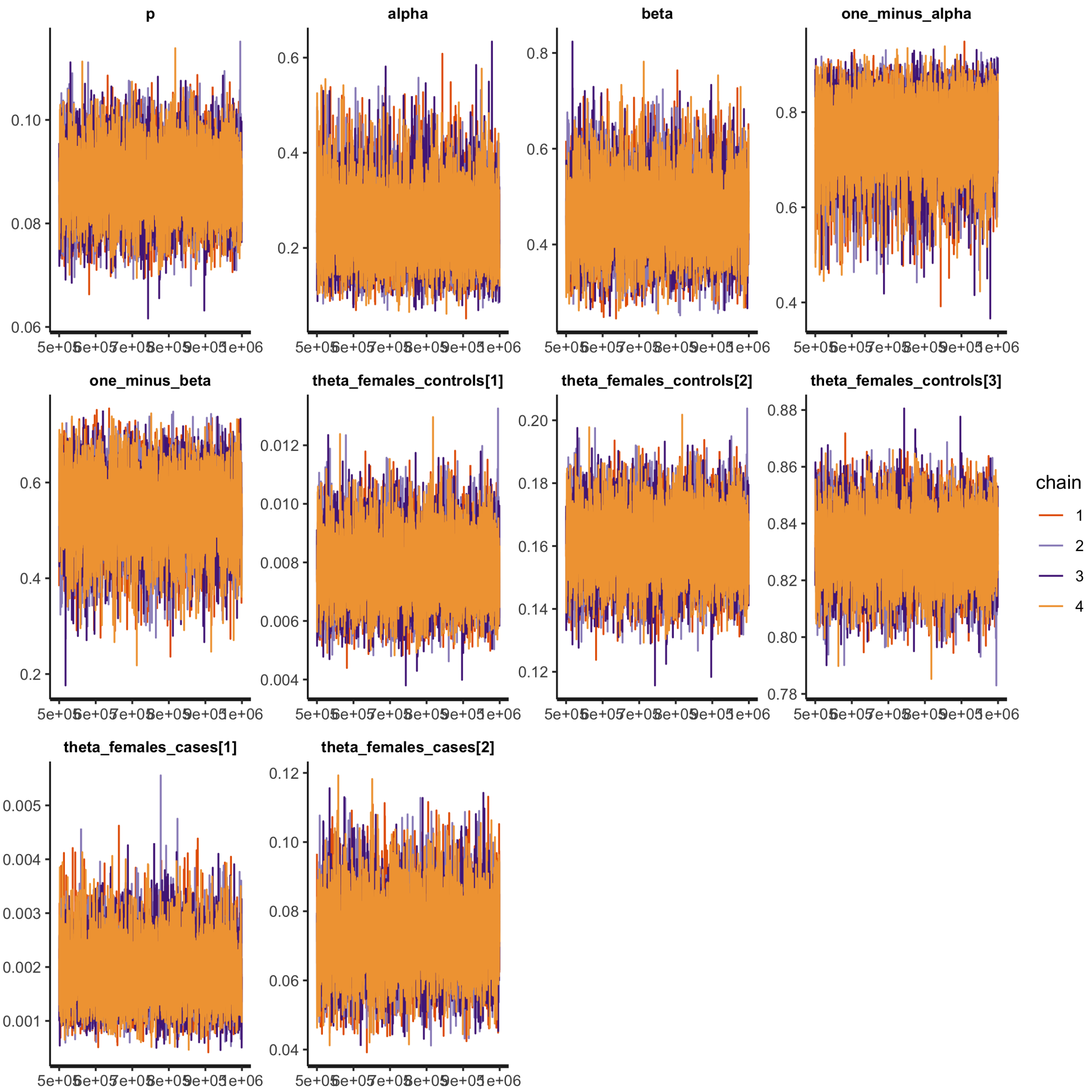


### Results

```
writeLines(sprintf('The allele frequency in the Pasthun ethnic group is %s (95%% CI %s-%s) (using Awab d
                   round(100*mean(thetas1$p), 1),
                   round(100*quantile(thetas1$p,probs = 0.025), 1),
                   round(100*quantile(thetas1$p,probs = 0.975), 1)))
```

```
## The allele frequency in the Pasthun ethnic group is 7.8 (95% CI 6.3-9.5) (using Awab data only)
```

```
writeLines(sprintf('The allele frequency in the Pasthun ethnic group is %s (95%% CI %s-%s) (meta-analysi
                   round(100*mean(thetas2$p), 1),
                   round(100*quantile(thetas2$p,probs = 0.025), 1),
```

```
                   round(100*quantile(thetas2$p,probs = 0.975), 1)))
```

```
## The allele frequency in the Pasthun ethnic group is 8.8 (95% CI 7.6-10.1) (meta-analysis)
```

```
writeLines(sprintf('The protective effect in hemi/homo-zygotes is %s%% (credible interval %s-%s) (using
                   round(100*(1-mean(thetas1$alpha))),
                   round(100*(1-quantile(thetas1$alpha,probs = 0.975))),
                   round(100*(1-quantile(thetas1$alpha,probs = 0.025)))))
```

```
## The protective effect in hemi/homo-zygotes is 66% (credible interval 39-85) (using Awab data only)
```

```
writeLines(sprintf('The protective effect in hemi/homo-zygotes is %s%% (credible interval %s-%s) (meta-a
                   round(100*(1-mean(thetas2$alpha))),
                   round(100*(1-quantile(thetas2$alpha,probs = 0.975))),
                   round(100*(1-quantile(thetas2$alpha,probs = 0.025)))))
```

```
## The protective effect in hemi/homo-zygotes is 75% (credible interval 58-88) (meta-analysis)
```

```
writeLines(sprintf('The protective effect in heterozygotes is %s%% (credible interval %s-%s) (using Awab
                   round(100*(1-mean(thetas1$beta))),
                   round(100*(1-quantile(thetas1$beta,probs = 0.975))),
                   round(100*(1-quantile(thetas1$beta,probs = 0.025)))))
```

```
## The protective effect in heterozygotes is 50% (credible interval 28-67) (using Awab data only)
```

```
writeLines(sprintf('The protective effect in heterozygotes is %s%% (credible interval %s-%s) (meta-analy
                   round(100*(1-mean(thetas2$beta))),
                   round(100*(1-quantile(thetas2$beta,probs = 0.975))),
                   round(100*(1-quantile(thetas2$beta,probs = 0.025)))))
```

```
## The protective effect in heterozygotes is 54% (credible interval 38-68) (meta-analysis)
```

```
writeLines(sprintf('The ratio of effect between hemi to hets is %s (credible interval %s to %s) (using A
                   round((mean((1-thetas1$beta)/(1-thetas1$alpha))),2),
                   round((quantile((1-thetas1$beta)/(1-thetas1$alpha),probs = 0.975)),2),
                   round((quantile((1-thetas1$beta)/(1-thetas1$alpha),probs = 0.025)),2)))
```

```
## The ratio of effect between hemi to hets is 0.78 (credible interval 1.3 to 0.43) (using Awab data onl
```

```
writeLines(sprintf('The protective effect in heterozygotes is %s (credible interval %s-%s) (meta-analysi
                   round((mean((1-thetas2$beta)/(1-thetas2$alpha))),2),
                   round((quantile((1-thetas2$beta)/(1-thetas2$alpha),probs = 0.975)),2),
                   round((quantile((1-thetas2$beta)/(1-thetas2$alpha),probs = 0.025)),2)))
```

```
## The protective effect in heterozygotes is 0.73 (credible interval 1-0.5) (meta-analysis)
```

```
writeLines(sprintf('The posterior probability that the effect is greater in hemi/homo-zygous deficients
                   round(mean(thetas1$beta > thetas1$alpha),2)))
```

```
## The posterior probability that the effect is greater in hemi/homo-zygous deficients is 0.88 (using Aw
```

```
writeLines(sprintf('The posterior probability that the effect is greater in hemi/homo-zygous deficients
                   round(mean(thetas2$beta > thetas2$alpha),2)))
```

```
## The posterior probability that the effect is greater in hemi/homo-zygous deficients is 0.98 (meta-ana
```

Figure 2 in paper

```
ind = names(mod_combined) %in% c('alpha','beta')
names(mod_combined)[ind] = c('hemi/homo-zygous','heterozygous')
```

```r
ind = names(mod_awab) %in% c('alpha','beta')
names(mod_awab)[ind] = c('hemi/homo-zygous','heterozygous')


x = 100 - 100*as.matrix(mod_combined, pars = c('hemi/homo-zygous','heterozygous'))
p1 = mcmc_intervals(x, point_est='median', prob_outer = 0.95) +
  scale_x_continuous(name = 'Reduction in prevalence of clinical vivax malaria (%)',
                     limits = c(0,100)) +
  theme(text=element_text(size=17),
        axis.line.y = element_blank(),
        axis.ticks.y = element_blank(),
        plot.title = element_text(size=34,hjust=0.5))# + ggtitle('Meta-analysis')
```

```
## Scale for 'x' is already present. Adding another scale for 'x', which will
## replace the existing scale.
```

```r
p1
```

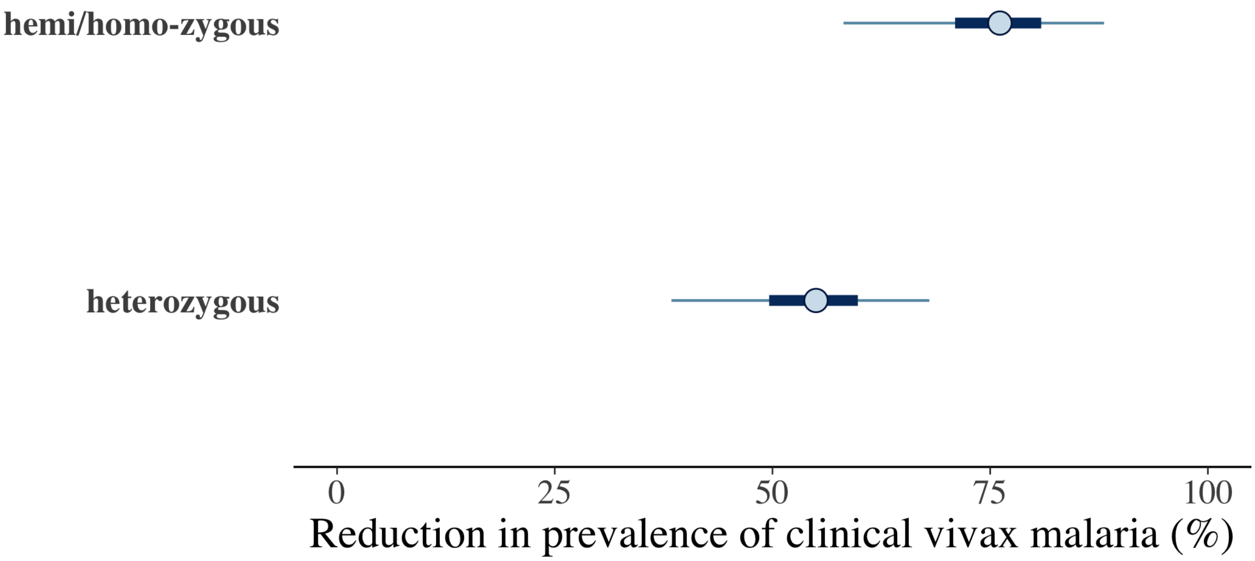


```r
x = as.matrix(mod_awab, pars = c('hemi/homo-zygous','heterozygous'))
p2 = mcmc_intervals(x) +
  scale_x_continuous(name = 'Proportion presenting with clinical vivax malaria relative to G6PD normals
                     limits = c(0,1))  +
  theme(axis.text=element_text(size=15),
        axis.line.y = element_blank(),
        axis.ticks.y = element_blank(),
        plot.title = element_text(size=24,hjust=0.5)) + ggtitle('Awab data only')
```

```
## Scale for 'x' is already present. Adding another scale for 'x', which will
## replace the existing scale.
```

```r
p2
```

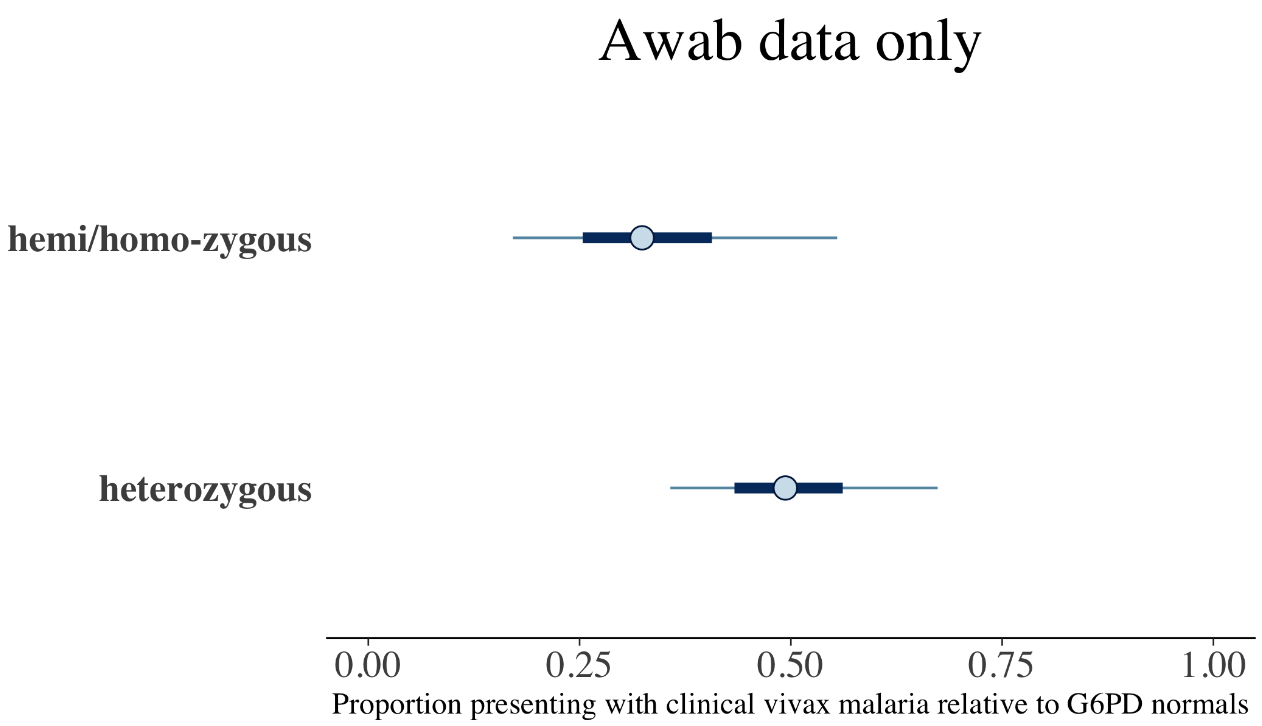


Make a plot for the paper with both results.

```
grid.arrange(p2, p1, nrow = 2)
```

Awab data only

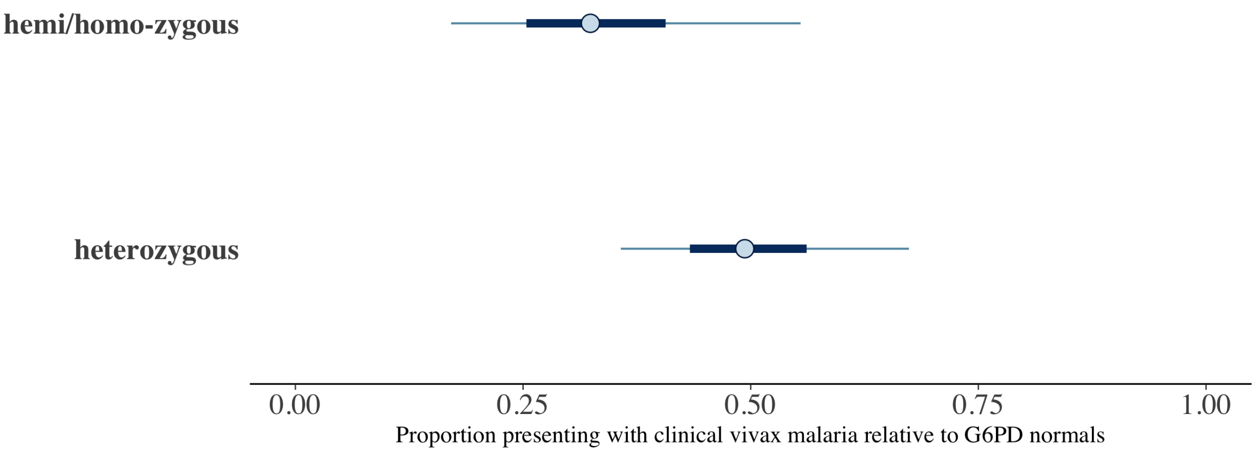


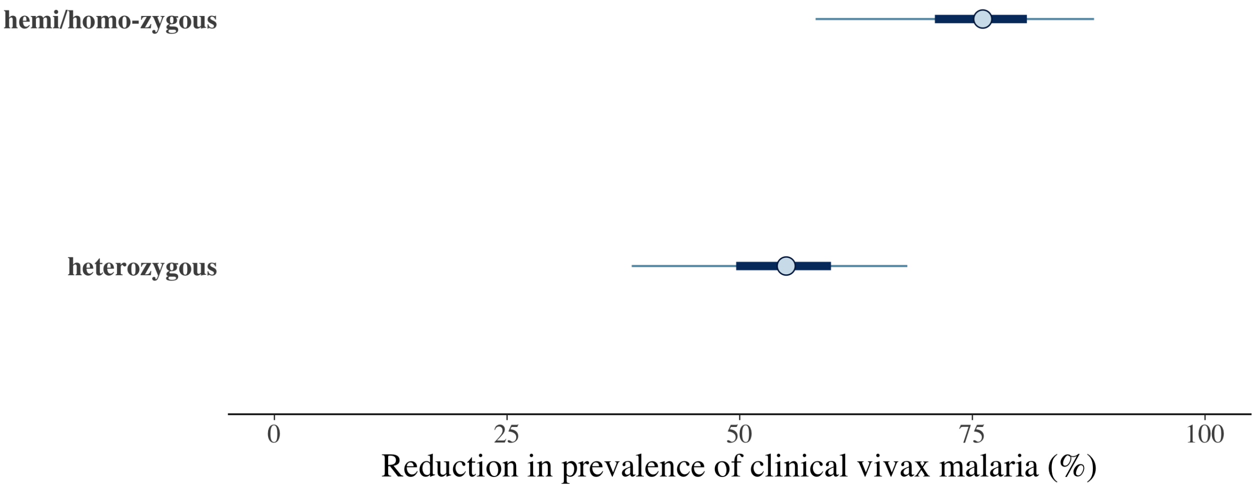
