## Supplementary figures and images for "Protective effect of Mediterranean type glucose-6-phosphate dehydrogenase deficiency against Plasmodium vivax malaria"

### Awab_results-1.png

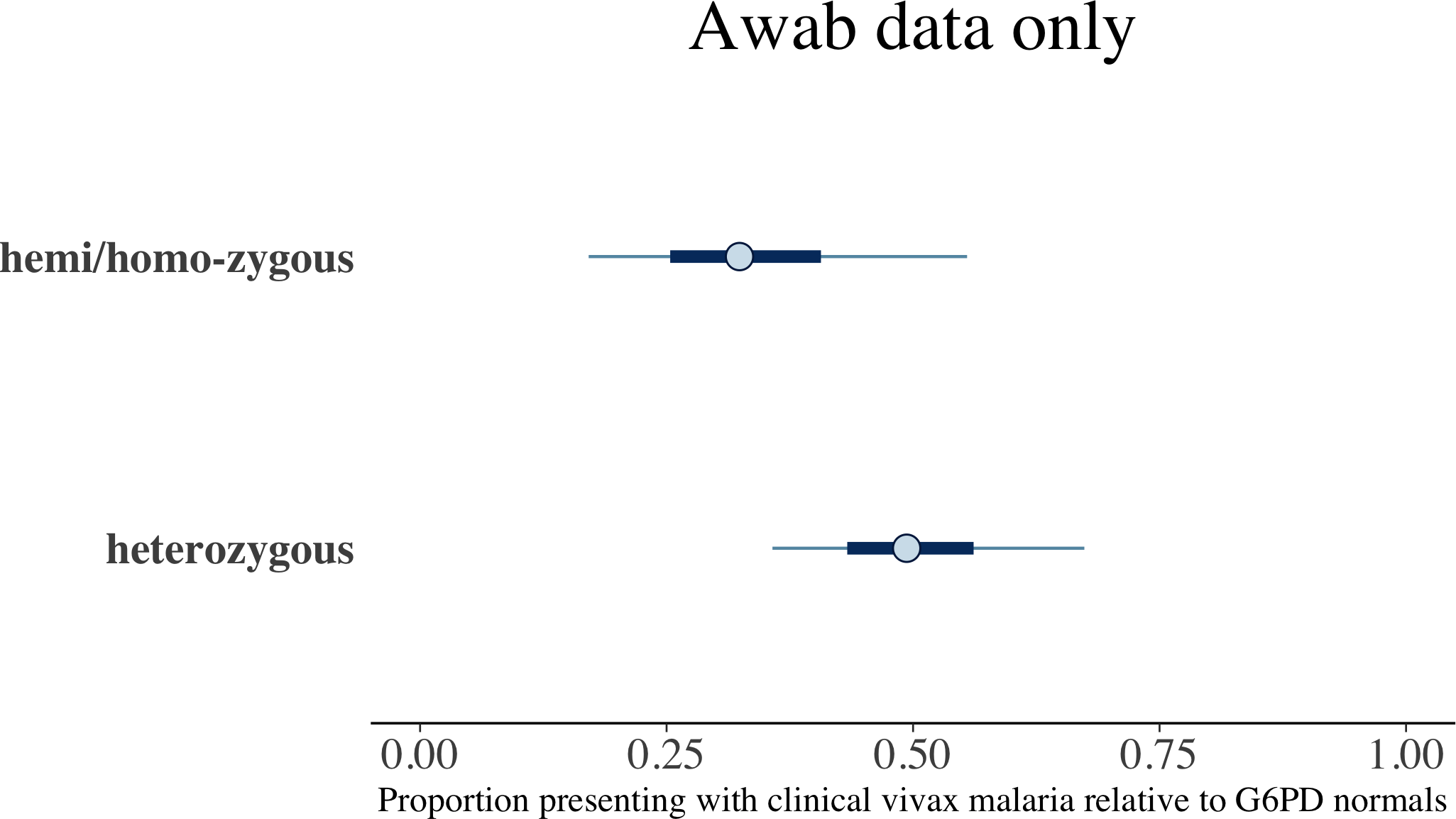

### meta_and_awab-1.png

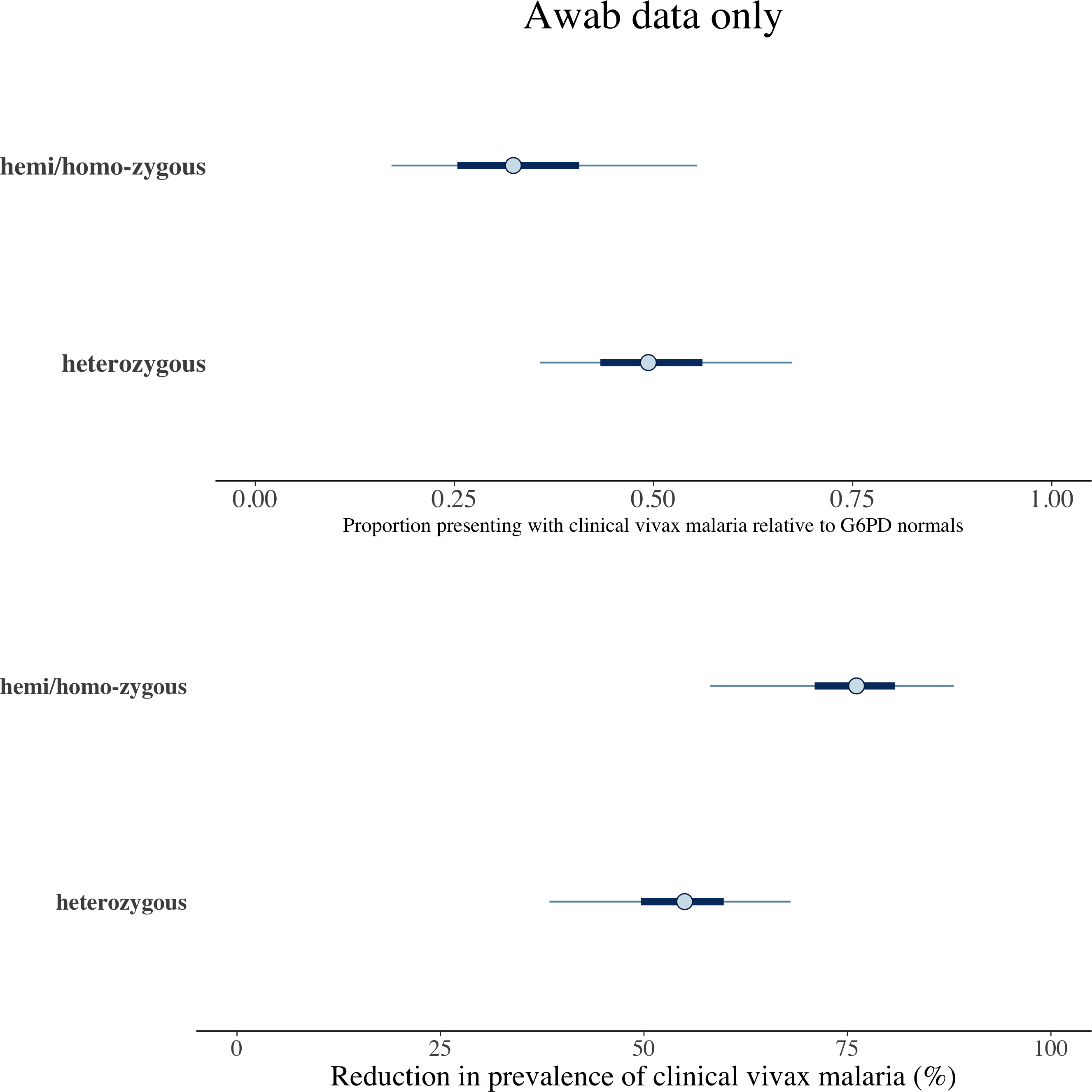

### metaanalysis_results-1.png

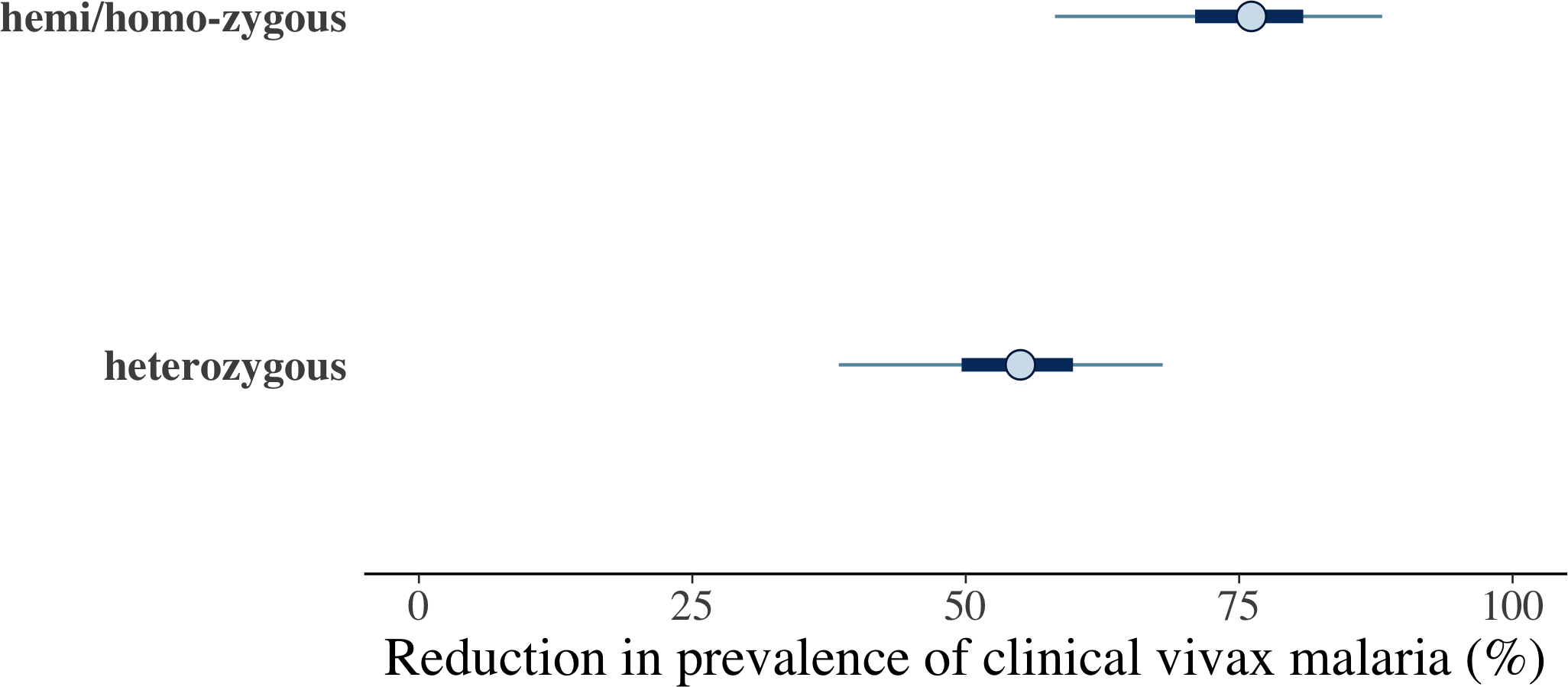

### unnamed-chunk-1-1.png

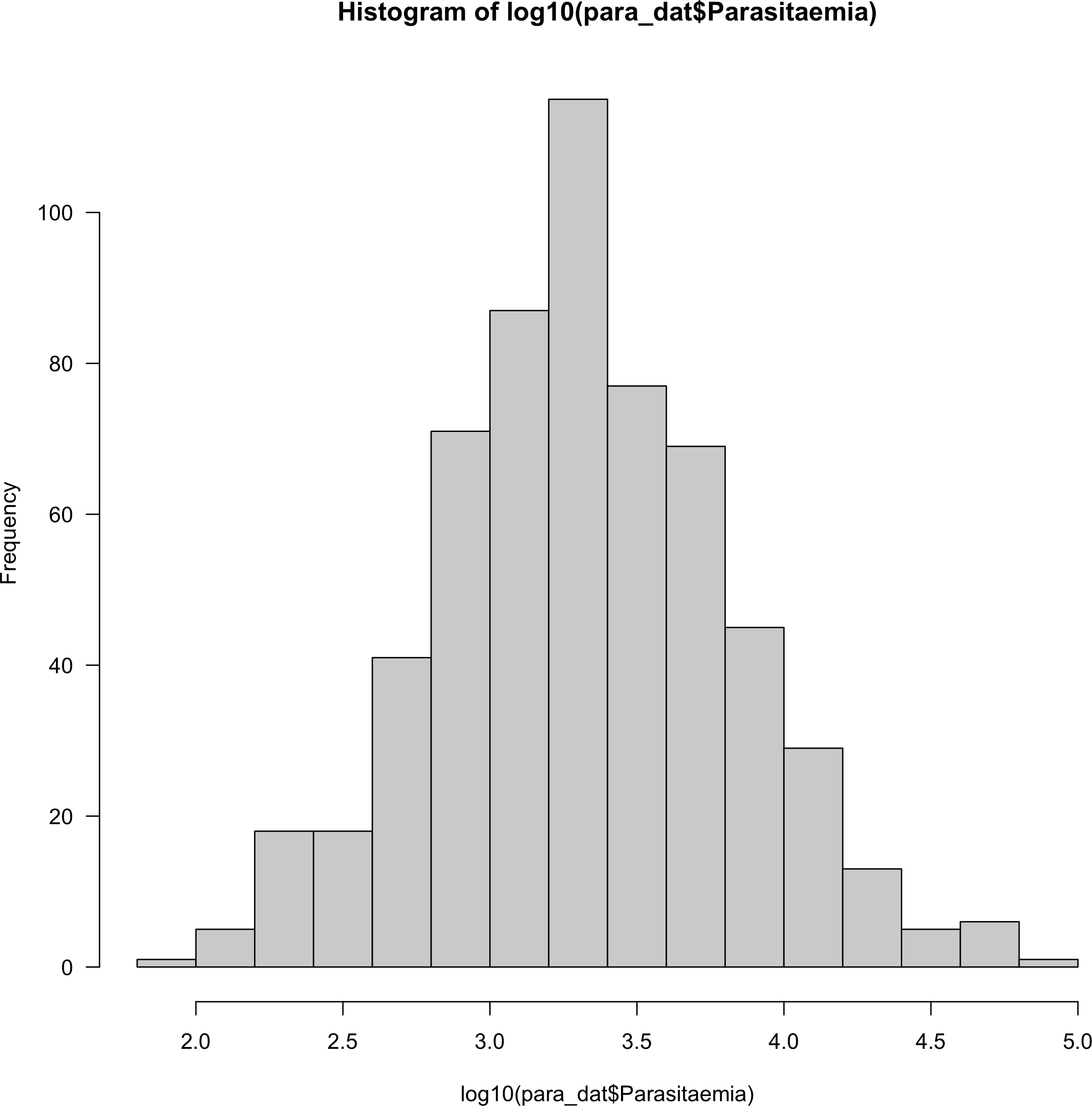

### unnamed-chunk-1-2.png

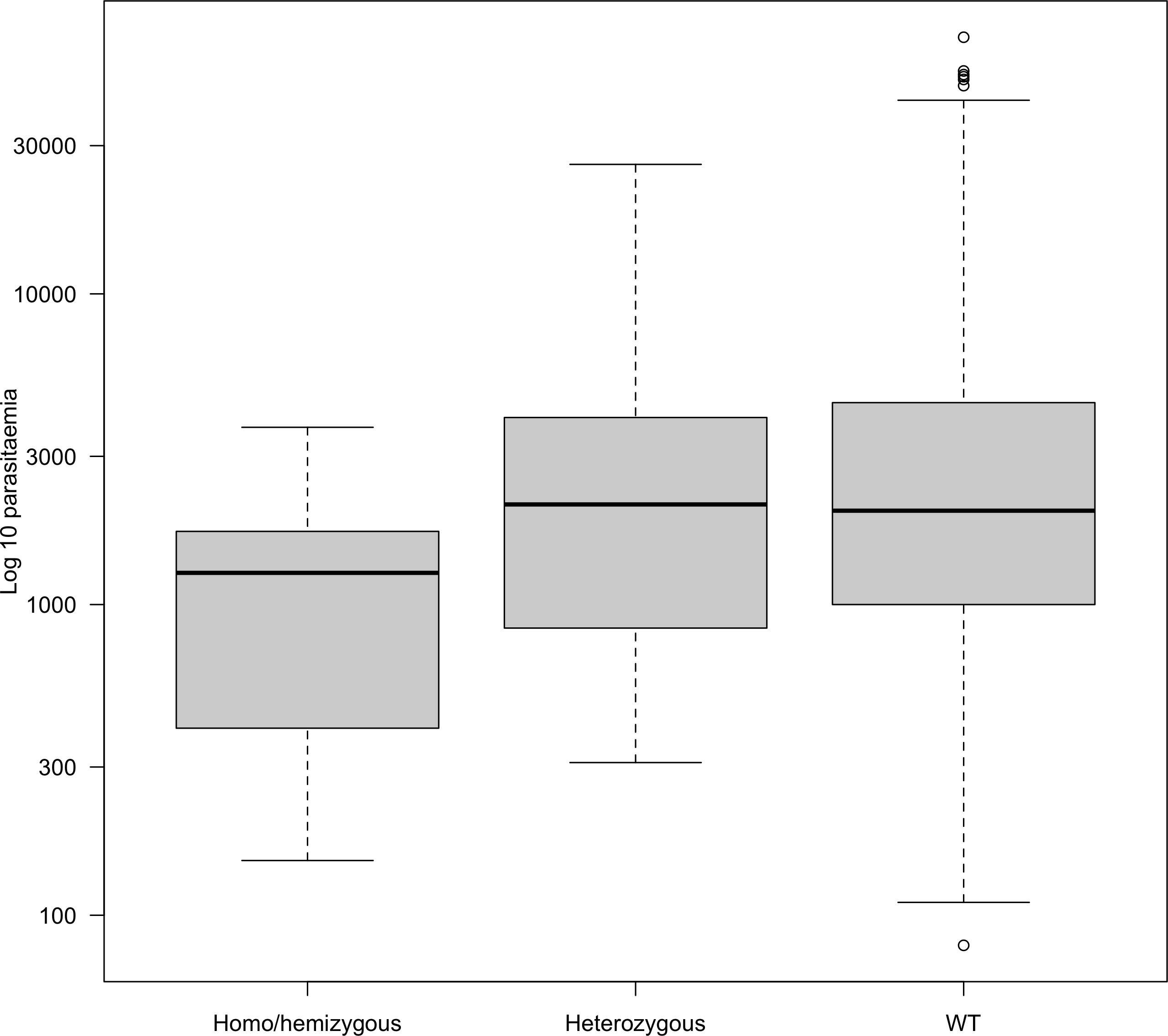

### unnamed-chunk-2-1.png

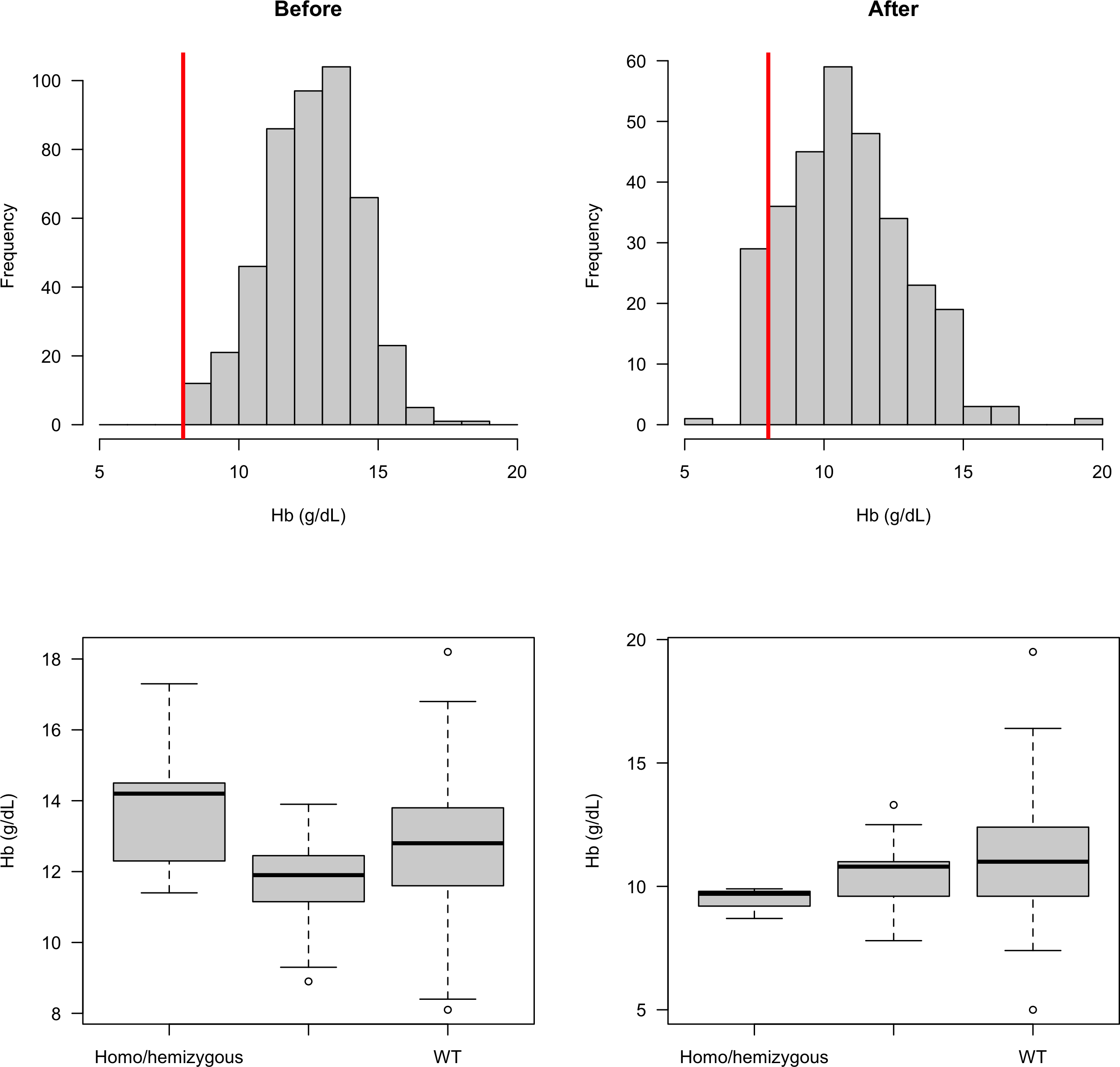

### unnamed-chunk-7-1.png

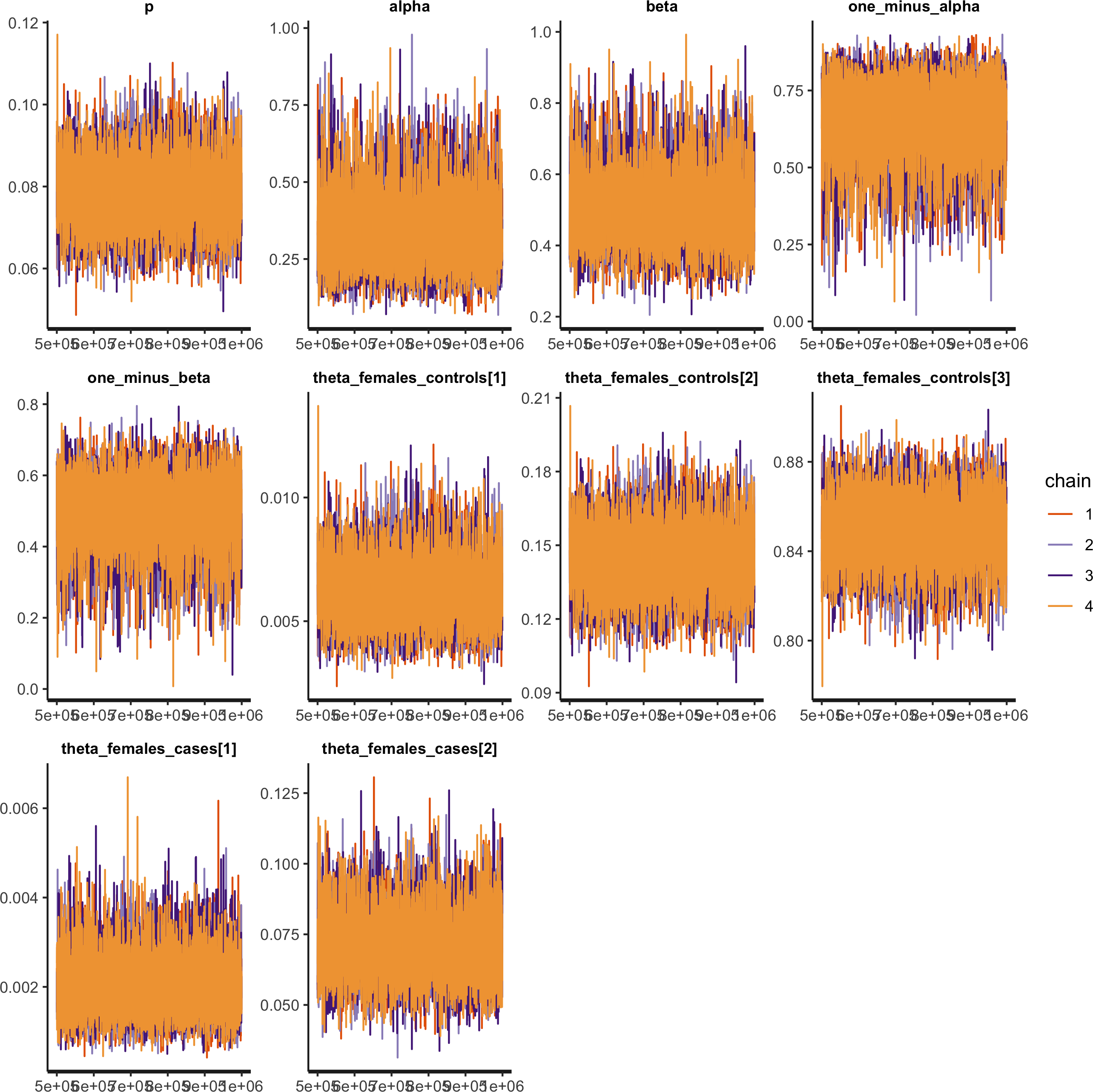

### unnamed-chunk-7-2.png

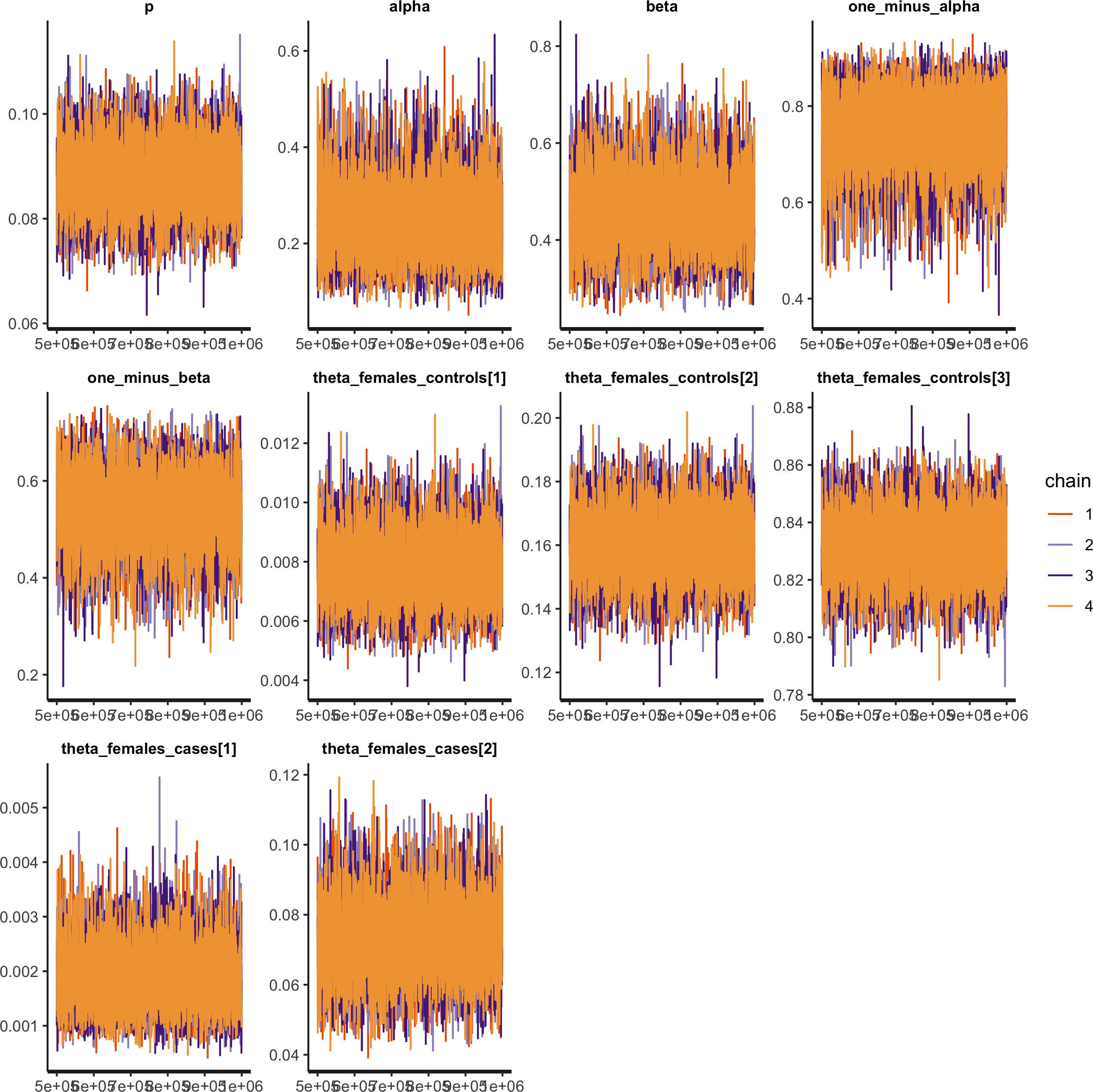
